# The impact of COVID-19 vaccination in the US: averted burden of SARS-COV-2-related cases, hospitalizations and deaths

**DOI:** 10.1101/2022.09.23.22280281

**Authors:** Teresa K. Yamana, Marta Galanti, Sen Pei, Manuela Di Fusco, Frederick J. Angulo, Mary M. Moran, Farid Khan, David L. Swerdlow, Jeffrey Shaman

## Abstract

By August 1, 2022, the SARS-CoV-2 virus had caused over 90 million cases of COVID-19 and one million deaths in the United States. Since December 2020, SARS-CoV-2 vaccines have been a key component of US pandemic response; however, the impacts of vaccination are not easily quantified. Here, we use a dynamic county-scale metapopulation model to estimate the number of cases, hospitalizations, and deaths averted due to vaccination during the first six months of vaccine availability. We estimate that COVID-19 vaccination was associated with over 8 million fewer confirmed cases, over 120 thousand fewer deaths, and 700 thousand fewer hospitalizations during the first six months of the campaign.

## Introduction

By August 1, 2022, SARS-CoV-2, the virus responsible for the COVID-19 pandemic, had caused over 90 million cases and 1 million deaths in the United States (1). While these numbers are likely affected by the widespread availability of SARS-CoV-2 vaccines, the precise impact of vaccination on the burden of COVID-19 disease is uncertain. Here we use a dynamic model, coupled with historical data, statistical inference methods, and hospitalization costs, to quantify the clinical and economic burdens of infections, hospitalizations, and deaths averted due to vaccination in the US, both cumulatively and in individual states, during the first approximately six months of vaccine availability when the wild type and alpha variants of SARS-CoV-2 were the predominant drivers of infection.

In mid-December 2020, the first SARS-CoV-2 vaccine received emergency use authorization in the US and was initially recommended for healthcare workers and long-term care facility residents, followed by adults aged 65 years and older, adults aged 16-64 with high-risk medical conditions and essential workers (2). By early April 2021, the vaccine recommendation was extended to the general population aged 16 years and older. Subsequent steps have seen recommended vaccine use for 12-15 year-olds (May 2021) and 5-11 year-olds (November 2021). Three different vaccines (two mRNA vaccines and one antiviral vector vaccine) with varying efficacy and estimates of duration of protection have been authorized for use in the US. However, vaccination delivery has been variable: it was initially limited by vaccine availability, with roughly 15 million doses provided in the first month, but reached a peak of roughly 90 million doses administered during April, 2021(1).

By early November 2021, 78% of the US population aged 12 years and older had received at least one dose of a SARS-CoV-2 vaccine, with heterogeneous distribution across age groups (97% of adults aged 65+ years vs 60% of persons aged 12-18 years) and across states (<65% in AL, ID, IN, LA, MS, ND, TN, WY, WV, compared with >90% in CT, MA, HI, VT, PA [2]). During the time period of vaccine rollout, variable levels of non-pharmaceutical interventions (NPIs), such as social distancing, closures of restaurants and bars, mask mandates and travel restrictions, were implemented across states with different start and end dates.

Here, we use a dynamic county-scale metapopulation model, previously used COVID-19 inference and projections (3-5), to conduct counterfactual simulations representing the effects of vaccination. These simulations are used to estimate the number of cases, hospitalizations, and deaths averted due to vaccination during the first six months of vaccine roll out.

## Methods

We used a metapopulation model with a Susceptible-Exposed-Infected-Recovered (SEIR) structure run at the county level, coupled with a data assimilation method (EAKF, the ensemble adjustment Kalman filter). We have previously used this framework for inference, forecasting and projections of influenza and SARS-CoV-2 infections at various locations and spatial scales (3-5). Here, we simulated SARS-CoV-2 transmission within and among the 3142 counties of the United States (see Supplementary Material).

Specifically, we first used the model-inference system to fit reported case counts in each county of the US [3] from the time of identification of the first COVID-19 cases in the United States in February 2020, through December 14, 2020, the date of first authorized SARS-CoV-2 vaccination in the US. The inferred values of parameters and state variables on December 14, 2020 served as initial conditions for the averted burden analysis.

We then included vaccination in the dynamical model structure using documented daily rates of vaccine administration (1, 6) (see Supplementary Materials). State-level daily vaccination data from the CDC COVID Data Tracker (1) were allocated proportionally to each county based on population size. Within each county, we assumed equal probability of vaccination regardless of prior infection status. We modeled the vaccine as producing direct effects only, with 90% effectiveness against infection (7-9) – i.e. 90% of vaccinated individuals with no prior immunity were fully protected while the remaining 10% receive no protection. Specifically, 90% of vaccinated individuals with no prior immunity were removed from the Susceptible pool and placed in the Recovered compartment 24 days after administration of the first dose. In the Recovered compartment, we did not distinguish between vaccinated individuals and individuals recovered from infection; given uncertainty and limited data on re-infections and waning, both were considered immune for the remainder of the simulation period. With the 24-day delay, the impact of vaccinations on the simulation begins on January 8th. This baseline scenario, retrospectively fitted to case counts, enabled estimation of the daily timeseries of epidemiological parameters, including R_t_, the time-varying reproductive number, for each county location from December 14, 2020 through June 3, 2021.

We ran the simulations through June 3, 2021 to focus on the impact of vaccination prior to the predominance of the Delta and Omicron variants (1). Given that the higher transmissibility and immune escape properties of the Delta and Omicron variants require substantial additional modifications of the dynamical model structure, as well as re-parametrization, we restricted our analysis to the December 14, 2020 through June 3, 2021, or the pre-Delta, time period, during which the vaccine provided strong protection against infection. To quantify the burden averted by vaccination, we compared the baseline vaccination scenario to 3 counterfactual no-vaccination scenarios simulated over the same time period. All counterfactual scenarios assumed no vaccinations (or, equivalently, 0% vaccine effectiveness) but varied transmissibility to mimic different levels of non-pharmaceutical intervention (NPI) response in the absence of vaccination:

- Counterfactual Scenario 1; A no-transmission-change, no-vaccination scenario in which the R_t_ daily time series for each location was as inferred for the baseline scenario;
- Counterfactual Scenario 2: A no-vaccination scenario in which R_t_ for each location-day was increased 10% with respect to the baseline scenario; and
- Counterfactual Scenario 3: A no-vaccination scenario in which R_t_ for each location-day was decreased 10% with respect to the baseline scenario.

These counterfactual scenarios represent potential population behaviors and policies that might have been effected in the absence of vaccination. Scenario 3 represents increased NPIs through policies and individual action; Scenario 2 represents a decrease of NPIs, perhaps due to pandemic fatigue. We compared cumulative SARS-COV-2 cases in the 3 no-vaccination scenarios to the baseline scenario at national and state levels, analyzed differences in averted cases among states, and identified factors correlating with vaccination success.

### Hospitalizations and Deaths

To calculate hospitalizations and deaths in the counterfactual scenarios, we made the assumption that excess cases would have continued to lead to hospitalizations and deaths at the same overall rate as they did in each state during the summer and fall of 2020, prior to vaccine availability. We applied a state-specific pre-vaccine Case Hospitalization Rate (CHR) and a Case Fatality Rate (CFR) multiplier to the total number of averted cases in each scenario. CHR was calculated as the number of COVID-19 hospitalizations divided by cases during August 1 – December 14, 2020. Hospitalization data were compiled from the HHS dataset (10) and cases from the Johns Hopkins Center for Systems Science and Engineering (JHU CSSE) COVID-19 data set (11, 12). August 2020 was the first full month with all states reporting daily COVID-19 hospitalizations. CFR was calculated as the number of COVID-19 deaths divided by the number of cases in each state during August 1 - December 14, 2020, using the JHU CSSE COVID-19 Data (11). We excluded deaths and cases prior to August 1, 2020, for consistency with the hospitalization data set, and because both the ascertainment rate (fraction of true infections that are reported as confirmed cases) and the infection fatality rate (fraction of true infections that resulted in death) were unstable during the initial wave of the pandemic (4).

### Hospitalization Costs

We calculated averted hospitalization costs by multiplying the distribution of estimated COVID-19 associated hospitalizations averted by the distribution of costs per hospitalization episode, obtained from the US-based Premier Healthcare COVID-19 claims database (13). The median (interquartile range Q1-Q3) cost per hospitalization episode was $12,046 ($6,309-$25,361).

## Results

### Initialization

At the start of the simulation period, December 14, 2020, it was estimated 74.1% (95% credible interval: 70.2 – 78.6) of the US population was susceptible, 0.8% (95% CrI 0.6-1.2%) exposed, 0.8% (95% CrI 0.6-1.0%) infectious and 24.3% (95% CrI 19.2-28.6%) recovered. Figure 1 shows the distribution of the estimated epidemiological parameters across states at the beginning of vaccine administration. The median estimated susceptible fraction, corresponding to the fraction of the population that had not yet been infected since the beginning of the pandemic varied by state and ranged from 58% (95% CrI 56%-61%) in North Dakota to 94% (93%-95%) in Vermont. The susceptible fraction was highest in northwestern and northeastern states. The time dependent reproductive number ranged from median 0.8 (0.7-1.4 95% CrI) in Minnesota to 2.0 (1.7-2.3 95% CrI) in Tennessee. The CFR prior to the start of vaccination varied from 0.5% in Alaska (95% CrI 0.3-0.7%) to 2.3% (95% CrI 2.1-2.6%) in Rhode Island. The CHR in the same period ranged from 3.8% (95% CrI 3.6-4.0%) in Alaska to 20.7% in Kentucky (95% CrI 20.5-20.9%). While there was a modest reduction in CHR at the national scale from the pre-vaccine period (8.8% August 1, 2020 – December 14, 2020) to the analysis period (7.8% December 15, 2020 – June 2, 2021), we did not observe consistent population level differences in CFR at the national level (1.5% during both time periods) nor to CHR and CFR at the state level.

**Figure 1.**
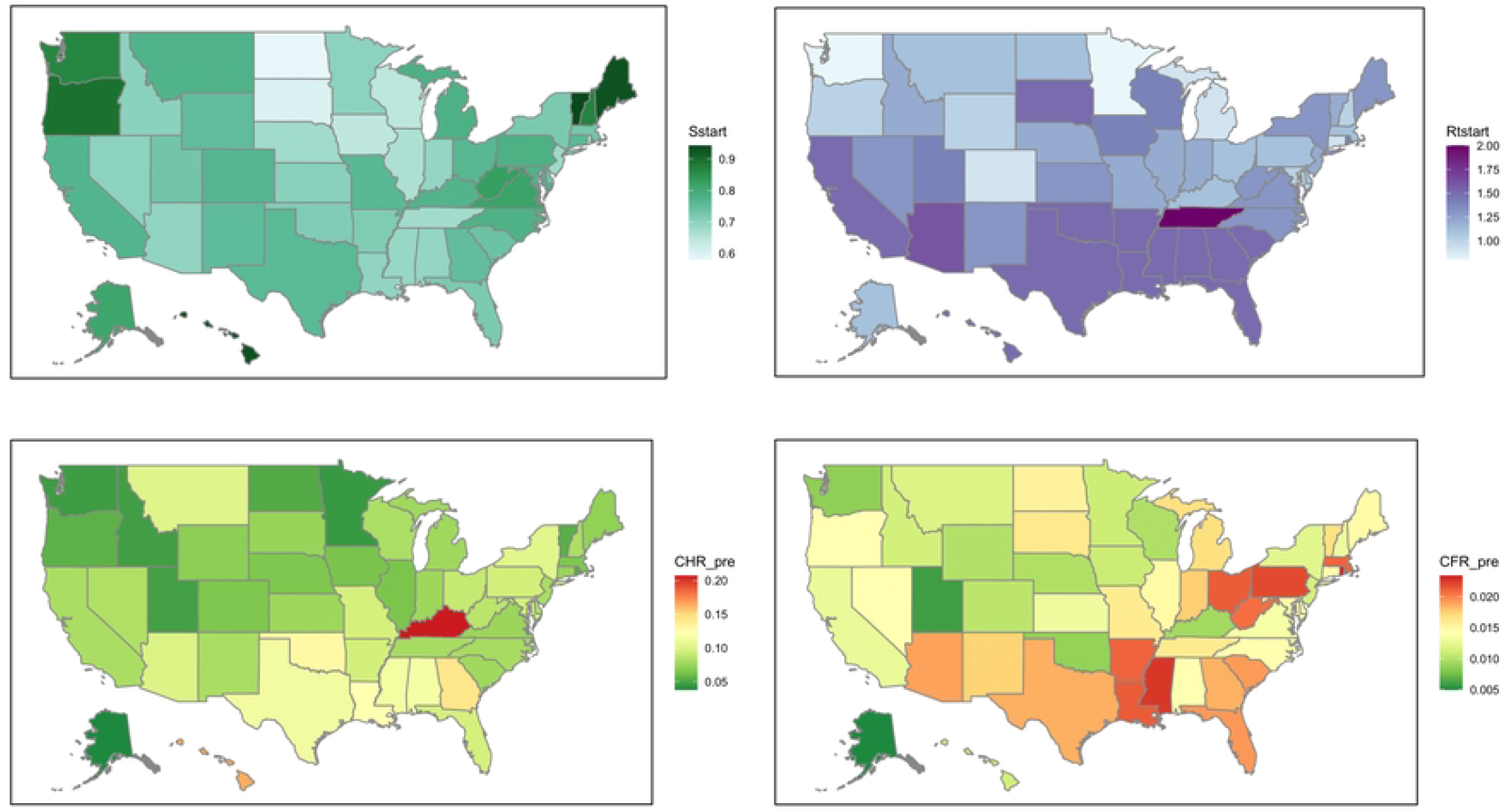
Upper Left: Population susceptibility, S (proportion of the population not yet infected), at the start of vaccine administration; Upper Right: Time-varying reproductive number, R_t_, at the start of vaccine administration; Lower Left: State-specific case hospitalization rate, CHR; Lower Right: State-specific case fatality rate, CFR. Color scales show the median values.

### Model results

Between December 14 and June 3, 2021, the baseline model estimated 16.1 million (95% CrI 15.1 – 18.3 million) total cases, 1.4 million (95% CrI 1.3 – 1.6 million) hospitalizations, and 246.7 thousand (95% CrI 230.4 – 279.6 thousand) deaths. These estimates were generally consistent with data reported by CDC COVID Data Tracker (1).

The time series of R_t_ resulting from fitting the baseline scenario from December 14, 2020 through June 4, 2021 is shown at the state and national level in Supplementary Figure S2. Note that R_t_ in this analysis refers to the time-varying basic reproductive number, not to be confused with the effective reproductive number R_eff_(t), which is R_t_ multiplied with the fractional susceptible population.

By June 4, 2021, 51% of the population in the US had received at least one dose of vaccine (1). Vaccine coverage differed widely by location, ranging from 35% of the population in Mississippi up to 74% in Vermont. The weekly number of vaccinations administered increased over time: initially at less than 5 million vaccinated per week but reaching a peak of 14 million vaccinated per week in April when vaccination was extended to the general population aged 16 and older (Figure S2).

Table 1 reports the cumulative averted COVID-19 cases, deaths, hospitalizations and hospitalization cost savings for the 3 scenarios, while Figure 2 shows the counterfactual scenarios and trends.

**Figure 2.**
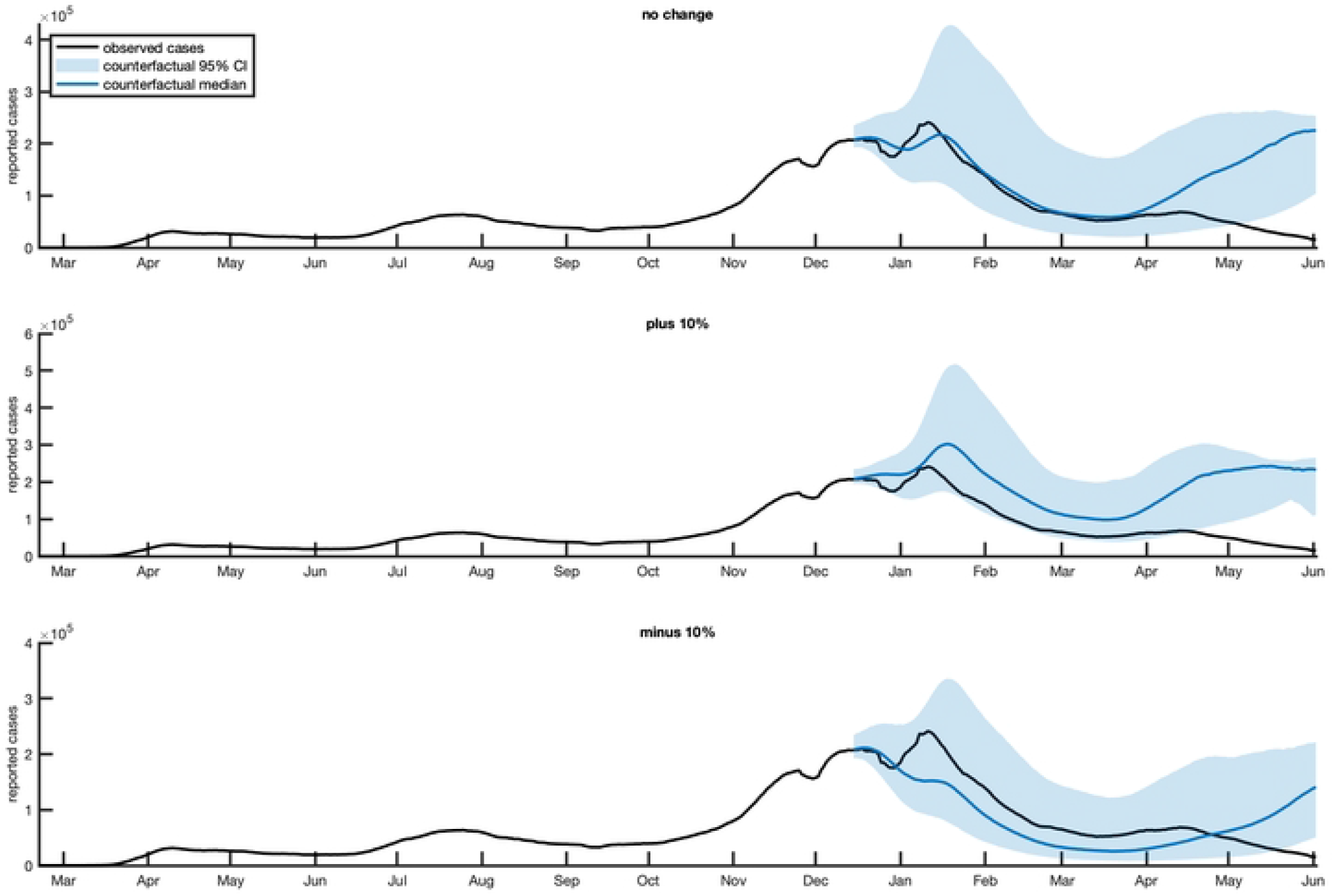
Modeled total COVID-19 Cases in Counterfactual Scenarios 1 (top panel), 2 (middle panel) and 3 (bottom panel) in the United States. The black line presents observed cases, the blue line indicates the median counterfactual projection, and the blue shaded area shows the 95% credible interval

**Table 1.** Total cumulative COVID-19 cases, deaths, hospitalizations and hospitalization cost savings

|  | Scenario 1<br>*No change in*<br>*transmission* | Scenario 2<br>*10% higher*<br>*transmission* | Scenario 3<br>*10% lower*<br>*transmission* |
| --- | --- | --- | --- |
| Cases averted<br>median, (95% CrI) | 8.1 million<br>(-4.8, 26.3 million) | 17.0 million<br>(1.5, 32.0 million) | -1.6 million<br>(-8.8, 18.2 million) |
| Deaths averted<br>median, (95% CrI) | 123.2 thousand<br>(-74.3, 403.0 thousand) | 260.1 thousand<br>(23.0, 489.7 thousand) | -25.1 thousand<br>(-134.8, 278.8 thousand) |
| Hospitalizations averted<br>median, (95% CrI) | 0.7 million<br>(-0.4, 2.3 million) | 1.5 million<br>(0.1, 2.8 million) | -0.1 million<br>(-0.8, 1.6 million) |
| Hospitalization cost<br>savings<br>median, (95% CrI) | \$7.0 billion<br>(\$-11.9, 112.0 billion) | \$17.3 billion<br>(\$0.9, 170.3 billion) | -\$0.9 billion<br>(-\$44.7, 70.1 billion) |

In the scenario with no change in transmission, we estimated that vaccination averted 8.1 million cases at the national level (median value, 95% CrI :[-4.8, 26.3] millions cases), 123.2 thousand deaths (median value, 95% CrI. :[-74.3, 403.0] thousand deaths) and 0.7 million hospitalizations (median value, 95% CrI :[-0.4, 2.3] millions hospitalizations). The median cost savings associated with averted hospitalization was $7.0 billion (median value, 95% CrI:[-$11.9, $112.0]) (see Table 1).

Increasing Rt by 10% with no vaccination in Counterfactual Scenario 2 roughly doubled the median cases averted nationally whereas decreasing Rt by 10% with no vaccination in Counterfactual Scenario 3 considerably reduced the averted burden during the approximately 6 months of analysis (Figure 2 and Table 1). In effect, the decreased Rt, representing increased NPIs, initially offsets the effects of no vaccination during the first 3 months when a more limited percentage of the population is effectively vaccinated. However, this effect decreases in mid-March as vaccination rates climb in the baseline scenario, and by May more cases are produced per day in Counterfactual Scenario 3 due to the absence of vaccination.

In all three counterfactual scenarios, the majority of averted cases occurred between April and June 2021 (Figure 2). For individual states, the median estimates of cases averted ranged from roughly1000 to 6400 cases per hundred thousand population (Figure 3). Median cumulative averted hospitalizations varied from 74 to 752 per hundred thousand and median cumulative averted deaths varied from 16 to 128 per hundred thousand. Higher averted case burden correlated with higher vaccination rate (R^2^=0.16) and higher population susceptibility at the beginning of the vaccination campaign (R^2^=0.21).

**Figure 3.**
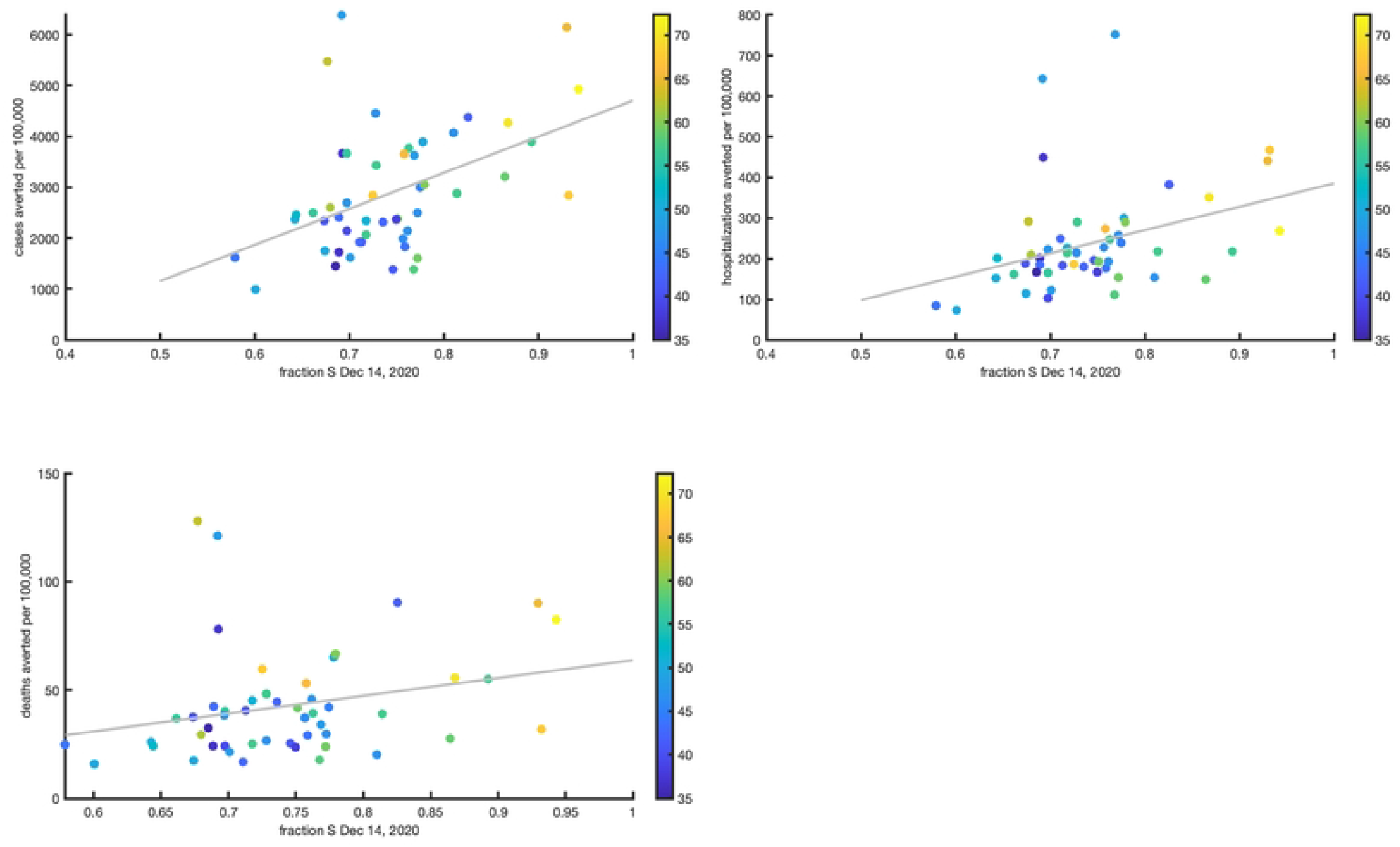
Total per capita averted cases (a), hospitalizations (b) and deaths (c) in each state between December 14, 2020 and June 2, 2021. The x-axis is the percent population vaccinated by June 2, 2021, and the y-axis is the averted cases/hospitalizations/deaths per 100,000 people. Each state is represented by a dot; the color scale of the dots indicate the estimated fraction of population susceptible at the beginning of vaccine rollout.

## Discussion

Evaluating the population-level impact of COVID-19 vaccination through mathematical modeling can provide useful insights to policy makers. Here, we leveraged a validated dynamical modeling approach, previously used for research and operationally to simulate county-level COVID-19 transmission, to quantify the additional burden of disease in alternate scenarios without vaccination. Our analyses show that under unchanged NPI levels, COVID-19 vaccination in the US cumulatively prevented 8.3 million cases, 681 thousand hospitalizations and 118 thousand deaths in the first 6 months of implementation. States with high vaccination coverage such as Maine averted as many as 6,000 cases per 100,000 individuals.

These simulations confirm findings from other modeling studies, set in the US and elsewhere, that have found a substantial impact of vaccination in terms of averted burden of disease. Shoukat et al found that vaccination was fundamental for reducing the spring/summer wave in NYC by reducing cases by one third and hospitalization and deaths by half during December 2020 through July 2021 (14). Vilches et al showed that vaccination may have averted more than 14 million cases, 241 thousand deaths and 1.1 million hospitalizations in the US by late June 2021(15). Moghadas et al. found an even stronger effect of vaccination with 26 million cases, 1.2 million hospitalizations and 279,000 deaths averted through the end of June 2021 (16). These three studies are in general agreement with our findings and indicate the US would have experienced a substantial wave of infections beginning in March/April 2021 in the absence of a vaccine (14-16). Large effects were found also in Israel by Haas et al. where two thirds of hospitalizations and deaths were averted with vaccination in the first four months of vaccine implementation (17).

Our study augments prior research in this field by providing further geographical granularity. The state-level analyses provide a dynamic picture revealing trends and differences in the public health response to the COVID-19 pandemic, which may be informative for state and local policymakers. Additionally, our study presents estimates of cost savings associated with vaccine-preventable severe disease (i.e. hospitalizations). It showed that the benefits of vaccination due to reduced hospitalization loads translated into cost-savings in the billions of dollars. Vaccination may also lessen other societal impacts associated with the pandemic (e.g. work productivity loss). The total economic impact may therefore be even greater than reported, and further studies elucidating those impacts are warranted.

Our study also adds to the existing literature by considering 3 counterfactual scenarios, all without vaccinations, but with varying R_t,_ that mimic different possible population responses to disease spread in the absence of a vaccine. The first counterfactual scenario is designed to quantify what the SARS-COV-2 related burden would have been without vaccinations if the population had maintained the same NPI measures as occurred with vaccination. The other two counterfactual scenarios are designed to explore the uncertainties of these estimates, as it is difficult to anticipate the public policy and population behavior response in the absence of a vaccine. Specifically, Counterfactual Scenario 2 represents a stronger relaxation of NPIs, possibly due to pandemic fatigue in the absence of an available vaccine, while Counterfactual Scenario 3 represents a reinforcement of NPIs during the 6 months of projections, assuming that the population would have responded with increased measures to control transmission. Counterfactual 3 shows that in the early months of the vaccine rollout, an increase in NPIs could have produced an even greater reduction of disease compared to vaccination as it occurred. However, while increased NPIs may have *slowed* transmission in the short term (the first months of vaccine rollout), those measures would not have been as effective as vaccination once the Alpha variant became established in the United States (Figure 2). The benefits of vaccination are seen in the difference between Counterfactual Scenario 3 and the baseline curve during the last month of simulation.

All 3 scenarios show that vaccination benefits were limited during the early months of vaccine rollout, and that most of the averted burden was realized in the last 2 months of the analyzed period. The winter peak of COVID-19 cases was reached in the US during mid-January 2021 just when the first vaccinations started to become effective. Vaccine availability constraints during the first months of the campaign restricted administration to portions of the population with increased risk of exposure and severe disease. It was not until April 2021 that vaccination was recommended for the general population aged ≥16 years. The combination of an initially slower rate of vaccination and a decreasing trend in transmission, with some states having a significant proportion of the population no longer susceptible to infection, narrowed the overall averted burden in the first months of 2021. Some exceptions occurred in states with larger initial susceptible fractions (e.g. Vermont); for these states the averted burden per hundred thousand was already significant in the early months of the vaccination campaign.

By March 2021, the Alpha variant, a SARS-CoV-2 strain with increased transmissibility relative to the wild type, became the predominant circulating serotype (1). This variant, combined with progressive relaxation of NPIs in most states, likely produced the increase of R_t_ inferred at this time. Simultaneously, the impact of vaccination, seen in the divergence between the baseline scenario and the no-vaccination scenario case curves, becomes much more evident at the national level (Figure 2).

A limitation of this analysis is that it relies on assumptions about whether and how the parameters inferred from the true observed course of the pandemic would have changed in the absence of a vaccine. Our primary counterfactual, Scenario 1, assumed that the parameters – including the disease transmission rates and the case ascertainment rate – would have been the same with or without a vaccine. We explored some of the sensitivity to this assumption by altering the time-varying reproductive number in Counterfactual Scenarios 2 and 3. However, these are very simplified representations, and one could just as well imagine dramatically different counterfactual scenarios.

We limit our analysis to a relatively short projection time: the first six months of the vaccination campaign. Our estimates are therefore not generalizable to the entire period of the vaccination campaign. In subsequent months, booster doses, the expansion of the Pfizer vaccine to children aged 5-11, waning immunity, and the establishment of the more virulent Delta and immune-evading Omicron variants have made estimation of vaccine effects more challenging. These later phases of the pandemic driven by new variants led to tens of millions of Covid-19 infections. Nevertheless, these early averted cases were crucial, as this period was prior to the widespread availability of antiviral medication, and with substantially lower population immunity against severe outcomes.

Additional assumptions should also be noted. The model structure is parsimonious and does not explicitly represent certain factors including population age structure, breakthrough infections or reinfections. We used a constant case hospitalization rate (CHR) and case fatality rate (CFR) for each state, computed based on COVID-19 outcomes during the 6 months before vaccination, to calculate counterfactual hospitalization and deaths in all scenarios. These choices ignore differences in age-specific behavior and probability of severe outcomes.

We also note that the full effect of COVID-19 vaccination on hospitalizations and deaths derives from two effects: those averted due to averted cases; and those averted due to improved outcomes in vaccinated individuals if infected. The estimates of averted hospitalizations and deaths in this analysis are restricted to the effect of averted cases and do not include reductions in the probability of hospitalization and death among the vaccinated if infected. Each of the approved COVID-19 vaccines has been shown to be highly effective in preventing severe outcomes in individuals infected by SARS-CoV-2. Here, we assumed that the contribution of the second effect was relatively small compared to the first, as the vaccines were shown to be highly effective at preventing infections in the short term after inoculation and against the strains circulating at the time of the study (18-20).

In conclusion, our analysis shows that COVID-19 vaccination reduced the burden of disease. Base case results indicate that COVID-19 vaccination was associated with over 8 million fewer confirmed cases, over 120 thousand fewer deaths, and 700 thousand fewer hospitalizations in the first six months of the campaign. As such, COVID-19 vaccines represented a critical component of the public health response to the COVID-19 pandemic in the US.

## Data Availability

Data will be held in a public repository at https://github.com/shaman-lab/

## Acknowledgements

This study was sponsored by Pfizer Inc. TKY, MG, SP and JS are employees of Columbia University, which received funding from Pfizer in connection with the development of this study and of this manuscript. JS and Columbia University disclose partial ownership of SK Analytics. JS discloses consulting for BNI. MDF, FJA, MMM, and FK are employees of Pfizer and may hold stock or stock options. DS was employed at Pfizer at the time this work was conducted and he may own stock or stock options.

